# Interrogating the causal effects of maternal circulating CRP on gestational duration and birth weight

**DOI:** 10.1101/2022.05.16.22275164

**Authors:** Jing Chen, Pol Sole Navais, Huan Xu, Christopher Flatley, Jonas Bacelis, Nagendra Monangi, Marian Kacerovsky, Mikko Hallman, Kari Teramo, Deborah A. Lawlor, Jeffrey C. Murray, Bo Jacobsson, Louis J. Muglia, Ge Zhang

## Abstract

**Background:** C-reactive protein (CRP) is an acute-phase marker of inflammation. Previous epidemiological studies have associated elevated maternal CRP levels during pregnancy with preterm birth risk and lower birth weight. However, the causal relationships behind these observed associations are not clear.

**Methods and Findings:** We utilized phenotype and genotype data on 10,734 mother/child pairs of European ancestry collected from six birth cohorts. We performed two-sample multivariate Mendelian Randomization (MR) using maternal non-transmitted alleles to interrogate the causal effect of maternal CRP on gestational duration and birth weight. Based on the non-transmitted alleles of 55 CRP associated single nucleotide polymorphisms (SNPs), a unit increase in log-transformed maternal CRP level was associated with a reduction of 1.58 days (95% CI: 0.23, 2.94, P = 0.022) in gestational duration and a reduction of 76.3g (95% CI: 18.2, 134.4, P = 0.010) in birth weight. The magnitude of the association between genetically increased CRP and birth weight was reduced after adjusting for gestational age. Utilizing the effect estimates of CRP-associated SNPs on birth weight from the UK Biobank genome-wide association (GWA) summary results, our results suggested one unit of genetically increased maternal CRP was associated with a decrease of 50.1g (95% CI: 24.7, 75.5, P = 9.6E-5) of birth weight (unadjusted by gestational age). We validated the results using a single genetic variant (rs2794520) near the CRP gene, which is less prone to horizontal pleiotropy bias.

**Conclusions:** Increased maternal CRP levels appear to causally reduce gestational duration and birth weight. The effect of maternal CRP on birth weight is partially due to its effect on gestational duration. The estimated causal effect sizes are consistent with previous epidemiological reports and this triangulation increases confidence in these results being causal.

## Introduction

Gestational duration and birth weight are highly correlated and canonical birth outcomes [1]. They are associated not only with perinatal morbidity and mortality but also with long-term adverse health outcomes [2,3]. Preterm birth (birth before 37 completed weeks of gestation) is the leading cause of mortality in children under five years [4,5]. Low birth weight, caused by shorter gestational duration and/or fetal growth restriction, is also associated with an increased risk of a broad range of short- and long-term health problems [6,7]. Inflammation is central to pregnancy, from implantation to parturition [8]. Disturbance of the normal inflammatory response due to endogenous or exogenous factors is believed to be a major mechanism behind multiple pregnancy complications (“the great obstetrical syndromes”), including preterm birth and fetal growth restriction [9].

C-reactive protein (CRP) is an acute-phase protein primarily synthesized in hepatocytes in response to infection, inflammation, and tissue damage [10]. Circulating CRP has been widely used as a diagnostic test for systemic inflammation in acute conditions and a marker of chronic inflammatory states. In healthy individuals, plasma CRP levels are usually below 3mg/L. Following an acute inflammatory stimulus such as bacterial infection, CRP concentration may increase over 10,000-fold [10]. Despite the sensitivity and wide range of CRP response to stimuli, baseline CRP concentrations within individuals are generally stable and slightly elevated CRP levels may indicate a state of low-grade inflammation in many complex diseases, including cardiovascular diseases [11], type 2 diabetes [12], cancers [13], and psychiatric disorders [14,15]. It has been speculated that in addition as a marker of inflammation, CRP might contribute to pathogenesis [10]. However, results from Mendelian randomization (MR) [16] studies did not support a causal role of elevated CRP in most of these diseases [17–20], except for some psychiatric disorders [21], with a notable protective effect on schizophrenia [22–24].

In obstetrics practice, maternal CRP concentration during pregnancy is used as a non-invasive marker of maternal systemic inflammatory response to intra-amniotic infection/inflammation, preterm premature rupture of membranes, urinary tract infection and pelvic inflammatory disease [25]. Pregnancy is a physiologic systemic inflammatory state and normal gestation is accompanied by an increase in the plasma concentrations of CRP from as early as 4 weeks to approximately 20 weeks of gestation [26,27]. Observationally, maternal inflammatory response is enhanced in many pregnancy complications [9], and increased levels of CRP during pregnancy have been observed in preterm birth [28–31], low birth weight [32,33], and pre-eclampsia [34,35]. It is unclear whether CRP is merely a marker of low-grade inflammation in these pregnancy complications, or if it plays an etiological role in the underlying pathological process. Resolution of this question will add knowledge of inflammatory mechanisms in pregnancy and may implicate new intervention strategies to prevent adverse pregnancy outcomes.

To interrogate this question, we applied a two-sample MR [36,37] to estimate the causal effects of maternal CRP on gestational duration and birth weight in over 10,000 mother/child pairs. Causal inference between maternal exposure and pregnancy outcomes is complicated by the maternally transmitted alleles. Conventional MR analysis using genetic instruments derived from maternal genotype cannot accurately estimate the causal effects of maternal exposure on pregnancy outcomes if the fetal genetic variants are not appropriately adjusted – association between maternal genetic instruments and offspring outcomes may arise from the fetal genetic effects of the transmitted alleles [38–40]. To overcome this problem, we adopted a haplotype-based MR approach that we have previously developed [41–43], in which the maternal non-transmitted and the paternal transmitted alleles are used respectively as valid instrumental variables (IVs) for maternal causal effect and fetal genetic effects. In addition, we sought to replicate our results in an independent sample by performing MR analysis on birth weight (not adjusted by gestational age) using the maternal and fetal genome-wide association (GWA) summary results from UK Biobank [44]. In this analysis, we used the weighted linear model (WLM) [45] to estimate the adjusted maternal and fetal effects to avoid confounding in causal inference due to genetic sharing between mothers and offspring.

## Materials and Methods

### Date sets

The study population consisted of 10,734 mother/child pairs from six birth studies (Table 1): these include three data sets collected from Nordic countries (The Finnish birth data set [FIN]) [46], The Norwegian Mother, Father and Child Cohort Study (MoBa) [47], and The Danish National Birth Cohort (DNBC) [48], a longitudinal birth cohort from the UK (the Avon Longitudinal Study of Parents and Children [ALSPAC]) [49,50], a preterm birth case/control cohort from the US (The Genomic and Proteomic Network for Preterm Birth Research [GPN]) [51], and the Hyperglycemia and Adverse Pregnancy Outcome study (HAPO) [52] with samples of European ancestry collected from the UK, Canada, and Australia. A detailed description of these data sets can be found in the Supplementary Text (S1 Text). Pregnancy-related phenotypes available included maternal height, age, pre-pregnancy BMI, gestational duration, and birth weight (except that birth weight was not available in the MoBa data set) (S1 Table). None of the cohorts had maternal measures of CRP during pregnancy. Maternal serum CRP levels measured in a follow-up 19 years after the pregnancy were available in ALSPAC [49,50]. This enabled us to examine whether the genetic instruments derived from a large GWA study in women and men [24] related similarly in women only.

**Table 1.** Birth data sets included in this study.

| Data set | Number of samples |  |  |  |
| --- | --- | --- | --- | --- |
|  | Pregnancies<sup>a</sup> | Mothers<sup>b</sup> | Children<sup>b</sup> | Duos<sup>c</sup> |
| FIN | 1371 | 1322 | 1217 | 1170 |
| MoBa | 1933 | 1804 | 1134 | 1009 |
| DNBC | 2038 | 1912 | 1865 | 1739 |
| HAPO | 1266 | 1203 | 1152 | 1089 |
| GPN | 440 | 419 | 364 | 343 |
| ALSPAC | 9806 | 7603 | 7587 | 5384 |
| Total | 16854 | 14263 | 13319 | **10734** |

a: Number of pregnancies with phenotype and genotype data
b: Number of individuals (mothers and infants) with phenotype and genotype data.
c: Number of mother/infant duos with phenotype and genotype data. In total, 10,734 mother/infant duos used in the analyses.
Abbreviations: ALSPAC, The Avon Longitudinal Study of Parents and Children; DNBC, The Danish National Birth Cohort; FIN, The Finnish birth data set; GPN, The Genomic and Proteomic Network for Preterm Birth Research; HAPO, The Hyperglycemia and Adverse Pregnancy Outcome Study; MoBa, The Mother Child data set of Norway.

This study involves reanalysis of existing data sets, and the analyses are consistent with the original consent agreements under which the genomic and phenotypic data were obtained. Therefore, additional ethics approval was not required.

### Genotype data

Individuals were genotyped using Affymetrix 6.0 (Affymetrix, Santa Clara, CA, USA) or various Illumina arrays (Illumina, San Diego, CA, USA). Similar genotype quality control procedures were applied across studies for all the data sets (S1 Text). Subjects of non-European ancestry were identified and excluded by principal components analysis (PCA). Genome-wide imputation was performed using Minimac3 [53] and the reference haplotypes extracted from phase 3 of the 1000 Genomes Project [54]. The phasing of SNP data was determined using Shapeit2 [55] in mother/child pairs. When phasing mother and child genotype data together, Shapeit2 accurately estimates maternally transmitted and non-transmitted, as well as paternally transmitted haplotypes – represented by h1, h2 and h3 respectively [43].

### Two-sample MR analysis in mother/child pairs

A two-sample MR analysis [36,37] was used to estimate the causal effect of CRP on gestational duration and birth weight (Figure 1). First, we chose the CRP-associated SNPs as the genetic instruments. These SNPs were extracted from a large GWA study of circulating concentration of CRP [24], which reported 58 genome-wide significant SNPs (P < 5E-8). Fifty-five of these CRP-associated SNPs were available in our data (S2 Table). According to the original study, these SNPs together explained about 6% CRP variance. The effects and standard errors of these SNPs on CRP were directly extracted from the original study [24], where the unit of the effect size was the natural log-transformed serum CRP level. This unit corresponds to a 2.72-fold change in CRP level, denoted as log(CRP) for simplicity.

**Figure 1.**
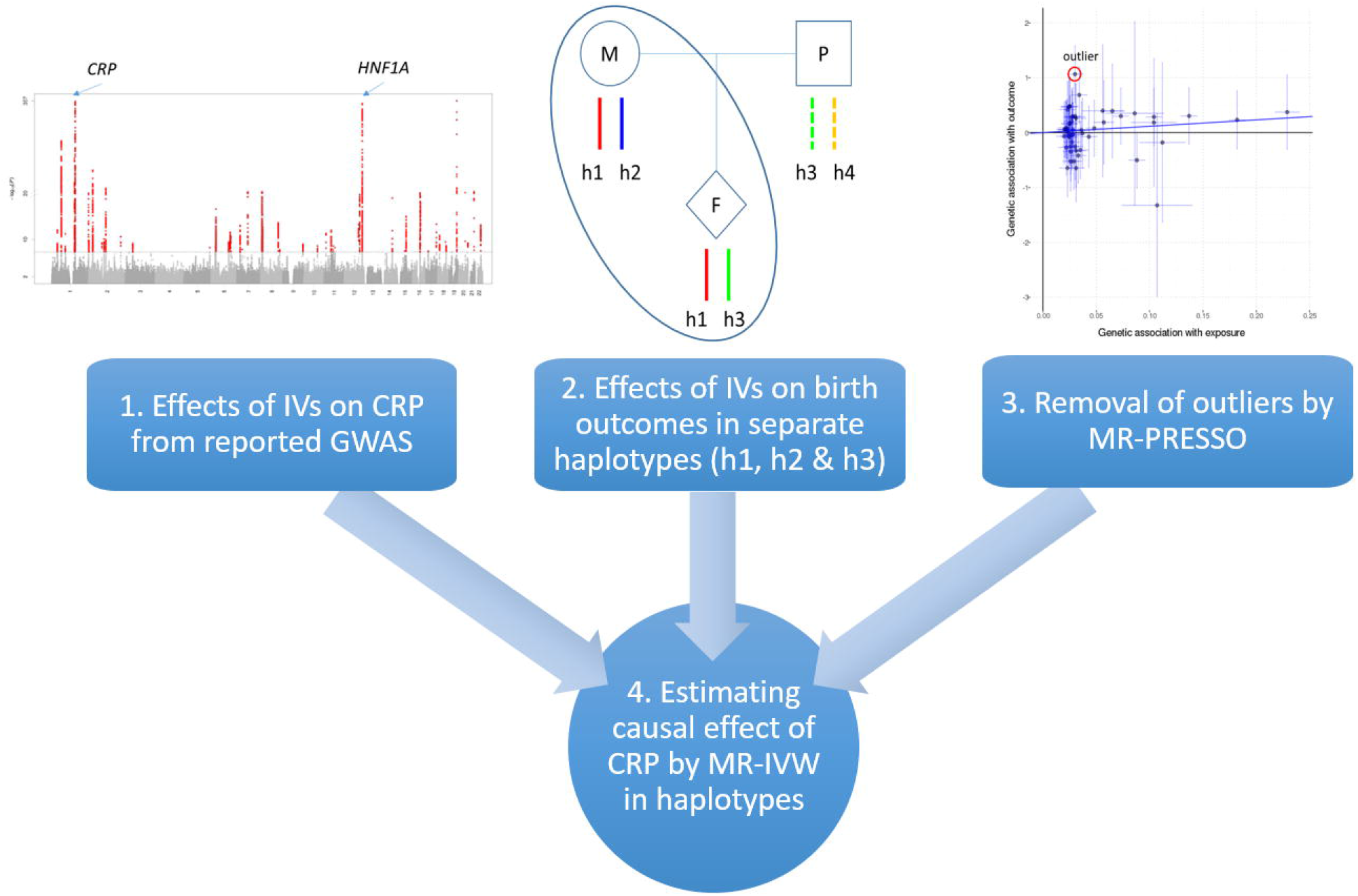
Workflow of the two-sample MR analysis. Abbreviations: IVs: instrumental variables; CRP: C-reactive protein; GWAS: genome-wide association study; MR: Mendelian Randomization; MR-PRESSO: Mendelian Randomization pleiotropy residual sum and outlier analyses; MR-IVW: Mendelian Randomization Inverse-Variance Weighted method.

In the second step, we estimated the effects of the SNPs on gestational duration and birth weight separately for the maternal transmitted alleles (h1), maternal non-transmitted alleles (h2), and paternal transmitted alleles (h3) using a haplotype-based association test previously described [42,43]. For each genetic variant, we fit a linear regression model: *y* ∼ *h*1 + *h*2 + *h*3 + covariates for continuous outcomes (gestational duration and birth weight); and a logistic regression model for preterm birth. The covariates for gestational duration and preterm birth included maternal age, maternal height, maternal BMI, and sex of the infants assigned at birth. For birth weight, gestational duration was also included as a covariate. The effects of the genetic variants were first estimated in each birth study separately and then combined through fixed-effect meta-analysis. The estimated effects of the maternal non-transmitted allele (h2) and the paternal transmitted allele (h3) were used to draw inferences about the maternal causal effect and the fetal genetic effect, respectively [42,43].

In the third step, we performed the MR pleiotropy residual sum and outlier analyses (MR-PRESSO) [56] to identify and remove outlying SNPs that might introduce bias due to horizontal pleiotropy. Specifically, the MR-PRESSO global test, the outlier test, and the distortion test were used to detect the presence of horizontal pleiotropy, the variants with a significant horizontal pleiotropic effect, and the distortion caused by the significant horizontal pleiotropic outlier variants respectively. Using this analysis, we were able to identify and remove the SNPs with significant horizontal pleiotropic effects from MR analysis to reduce potential bias in causal inference.

Lastly, we employed the Inverse-Variance Weighted (IVW) method [57] to estimate the causal effect of CRP on a pregnancy outcome by fitting a regression line between the reported effects of the IVs (i.e. CRP-associated SNPs) on CRP and the estimated effects of these IVs on a outcome, weighted by the inverse variances of their effects on the outcome. The intercept of the line was forced to zero, and the slope was then the estimated causal effect of the exposure on the outcome. The maternal effect and the fetal genetic effect of genetically elevated CRP on pregnancy outcomes were assessed using the maternal non-transmitted alleles (h2) and the paternal transmitted alleles (h3) respectively.

We also performed single IV MR using rs2794520 – the SNP with the strongest association with CRP levels. This SNP is a *cis*-protein quantitative trait locus (∼3kb downstream of the CRP gene), which is less prone to horizontal pleiotropy bias. In the original publication [24], this SNP explained up to 1.4% of CRP variance by itself. The Wald ratio method [58] was used to estimate the effects of CRP on pregnancy outcomes using this SNP as a single IV.

### Two-sample MR analysis using UK Biobank summary results

To replicate our findings in an independent sample by a different MR approach using an alternative method to control for fetal genotype, we performed a two-sample MR analysis to study the relationship between CRP and birth weight (unadjusted for gestational duration), using the UK Biobank GWA summary results. The effects of the CRP-associated SNPs on birth weight were extracted from the MR-Base database [59], and the MR analyses were performed using the R package TwoSampleMR. The results ‘UKB-a:318’ and ‘UKB-a:198’ in the MR-Base were the UK Biobank GWA summary results for ‘birth weight of the first child’ and ‘birth weight’ that were processed by the Neale Lab in 2017 (http://www.nealelab.is/uk-biobank). The former is an approximation of the maternal effects on birth weight confounded by the fetal effects of the transmitted alleles, whereas the latter is an approximation of the fetal effects confounded by the maternal effects of the maternally transmitted alleles. Therefore, we adjusted the former for fetal genotype and the latter for maternal genotype using the weighted linear model (WLM) adjusted analyses [45] to derive unbiased estimates of maternal and fetal effects. Briefly, the adjusted maternal effect 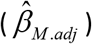 and fetal effect 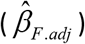 were calculated from the GWA summary results using maternal 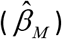 and fetal genotypes 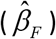.

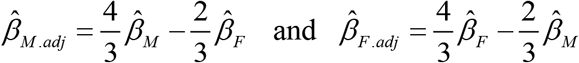

The adjusted maternal effect 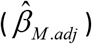 and fetal effect 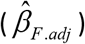 of the CRP-associated SNPs on birth weight were then used to estimate the maternal causal effect and fetal genetic effect of CRP on birth weight (unadjusted by gestational duration) using the Two-sample MR analysis as described above.

## Results

### Effects of CRP on gestational duration and birth weight in mother/child pairs

First, we attempted to validate the CRP-associated SNPs that we used as genetic instruments for circulating maternal CRP concentrations. In our data sets, CRP concentrations were not measured during pregnancy; however, the CRP concentrations of the mothers were measured 19 years after the pregnancy in the ALSPAC study. Overall, the 55 CRP-associated SNPs together and the SNP rs2794520 alone could explain 5.7% and 1.4% of the log-transformed CRP variance, respectively, which were similar to the reported numbers in the original publication [24].

The effects of the 55 CRP-associated SNPs on gestational duration, preterm birth, and birth weight (with or without adjustment for gestational duration) were estimated through the haplotype-based association tests in each data set separately. The estimated effects of these SNPs were then combined using fixed-effect meta-analysis (S3 Table).

MR-PRESSO analysis was used to identify outlier SNPs with significant horizontal pleiotropic effects. We detected no outliers for the maternal non-transmitted alleles (h2) but one outlier for the paternal transmitted allele (h3) in the analysis of gestational duration and it was removed from the MR analysis accordingly (S4 Table). Using the maternal non-transmitted alleles (h2) of the 55 CRP associated SNPs, the estimated effect of maternal CRP on gestational duration was −1.58day/log(CRP) (95% CI: −2.94, −0.23, P = 0.022) (Table 2, Figure 2 and S1 Figure), i.e., every unit of genetically elevated log-transformed maternal CRP level was associated with 1.58 days shorter gestational duration. Using the non-transmitted allele of rs2794520 as the single IV, the Wald ratio estimated effect size was - 2.75day/log(CRP) (95% CI: −5.65, 0.15; P = 0.063). When gestational duration was analyzed as a binary outcome (i.e. preterm vs term birth), the estimated effect of genetically elevated maternal CRP on preterm birth risk had an OR = 1.25 (95% CI: 0.90, 1.75), which was directionally consistent with findings for gestational duration, however, it was not statistically significant (P = 0.20). Similarly, we also observed significant causal associations between maternal CRP and unadjusted birth weight in both multiple IVs or single IV analyses (Table 2, Figure 3 and S2 Figure), with estimated effects of - 76.3g/log(CRP) (95% CI: −134.4, −18.2; P = 0.01) and −136.9g/log(CRP) (95% CI: −261.5, −12.2; P = 0.03) respectively. After adjusting for gestational duration, the estimated causal effects based on maternal non-transmitted alleles (h2) were reduced and no longer significant (Table 2, Figure 3 and S3 Figure).

**Table 2.** Haplotype-based MR analysis of CRP on pregnancy outcomes.

| Outcome<sup>a</sup> | Haplotype<sup>d</sup> | MR based on 55 CRP SNPs |  |  |  | MR based on rs2794520 |  |  |  |
| --- | --- | --- | --- | --- | --- | --- | --- | --- | --- |
|  |  | Estimate | Std Err | 95% CI | P-value | Estimate | Std Err | 95% CI | P-value |
| Gestational duration | h1 | -1.20 | 0.72 | -2.62, 0.23 | 0.099 | -1.18 | 1.49 | -4.10, 1.73 | 0.426 |
|  | h2 | -1.58 | 0.69 | -2.94, -0.23 | 0.022 | -2.75 | 1.48 | -5.65, 0.15 | 0.063 |
|  | h3 | 1.05 | 0.76 | -0.44, 2.54 | 0.166 | 1.27 | 1.47 | -1.62, 4.16 | 0.388 |
| Preterm birth | h1 | 0.13 | 0.19 | -0.24, 0.51 | 0.482 | 0.6 | 0.38 | -0.14, 1.34 | 0.110 |
|  | h2 | 0.22 | 0.17 | -0.11, 0.56 | 0.197 | 0.54 | 0.37 | -0.19, 1.26 | 0.147 |
|  | h3 | 0.09 | 0.18 | -0.26, 0.44 | 0.621 | 0.44 | 0.37 | -0.29, 1.16 | 0.238 |
| Birth weight (unadjusted)<sup>b</sup> | h1 | -36.7 | 36.9 | -109.1, 35.7 | 0.320 | 8.3 | 64.3 | -117.7, 134.3 | 0.897 |
|  | h2 | -76.3 | 29.6 | -134.4, -18.2 | 0.010 | -136.9 | 63.6 | -261.5, -12.2 | 0.031 |
|  | h3 | -0.5 | 31.4 | -61.9, 61.0 | 0.988 | -67.2 | 63.6 | -191.8, 57.3 | 0.290 |
| Birth weight (adjusted)<sup>c</sup> | h1 | -12.9 | 24.09 | -60.1, 34.3 | 0.592 | 34.2 | 48.0 | -59.8, 128.2 | 0.476 |
|  | h2 | -32.9 | 24.2 | -80.3, 14.6 | 0.175 | -75.7 | 47.5 | -168.7, 17.4 | 0.111 |
|  | h3 | -28.0 | 22.0 | -71.2, 15.2 | 0.204 | -90.1 | 47.6 | -183.3, 3.1 | 0.058 |

a: The units of effect size estimates are day (gestational duration), log odds (preterm birth) and gram (birth weight).
b: MR analysis of birth weight without including gestational duration as a covariate
c: MR analysis of adjusted birth weight by including gestational duration as a covariate
d: MR analysis based on maternal transmitted (h1), maternal non-transmitted (h2), and paternal transmitted alleles (h3)

**Figure 2.**
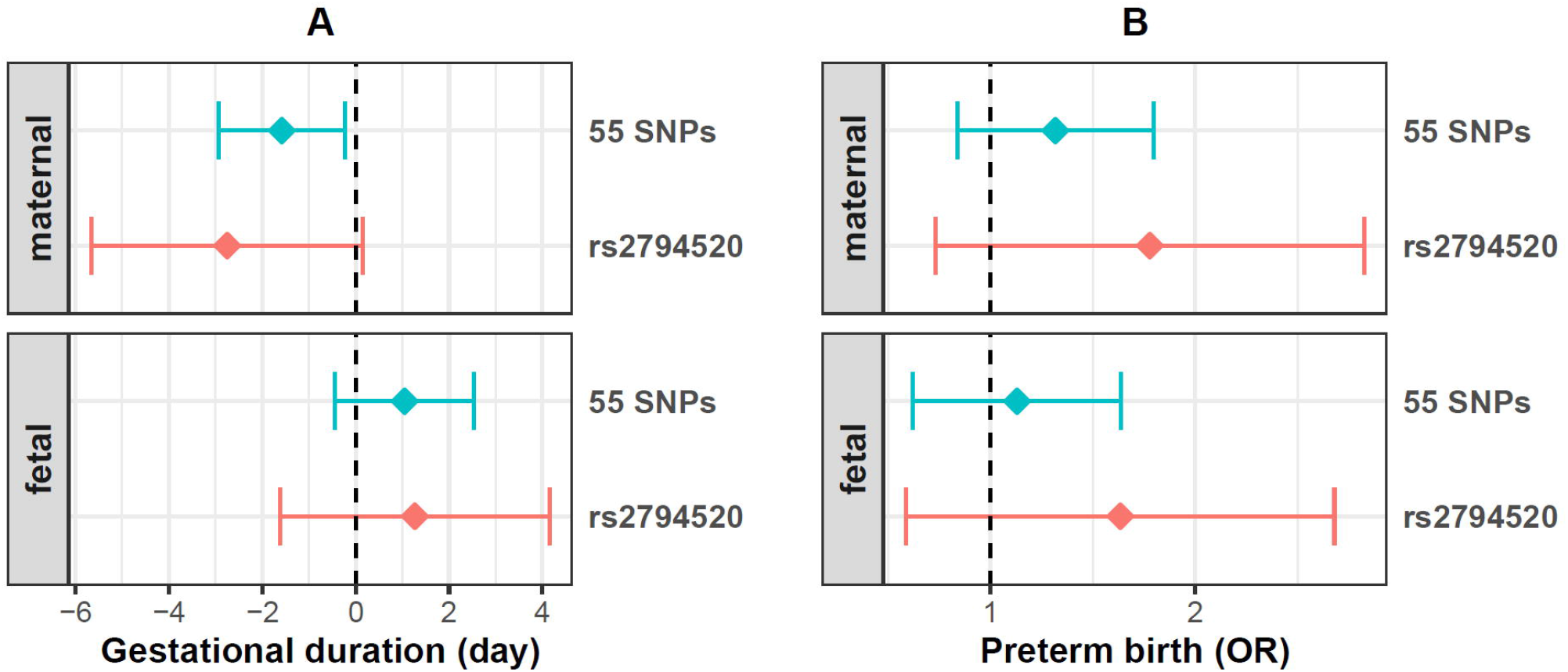
Estimated maternal effects and fetal genetic effects per one unit genetically increased log-transformed CRP levels on gestational duration (A) and preterm birth risk (B). The maternal effects were estimated by maternal non-transmitted alleles (h2) and the fetal genetic effects were estimated by paternal transmitted alleles (h3). The estimates based on multiple (55 SNPs, blue) or single (rs2794520, red) genetic variant(s) are shown.

**Figure 3.**
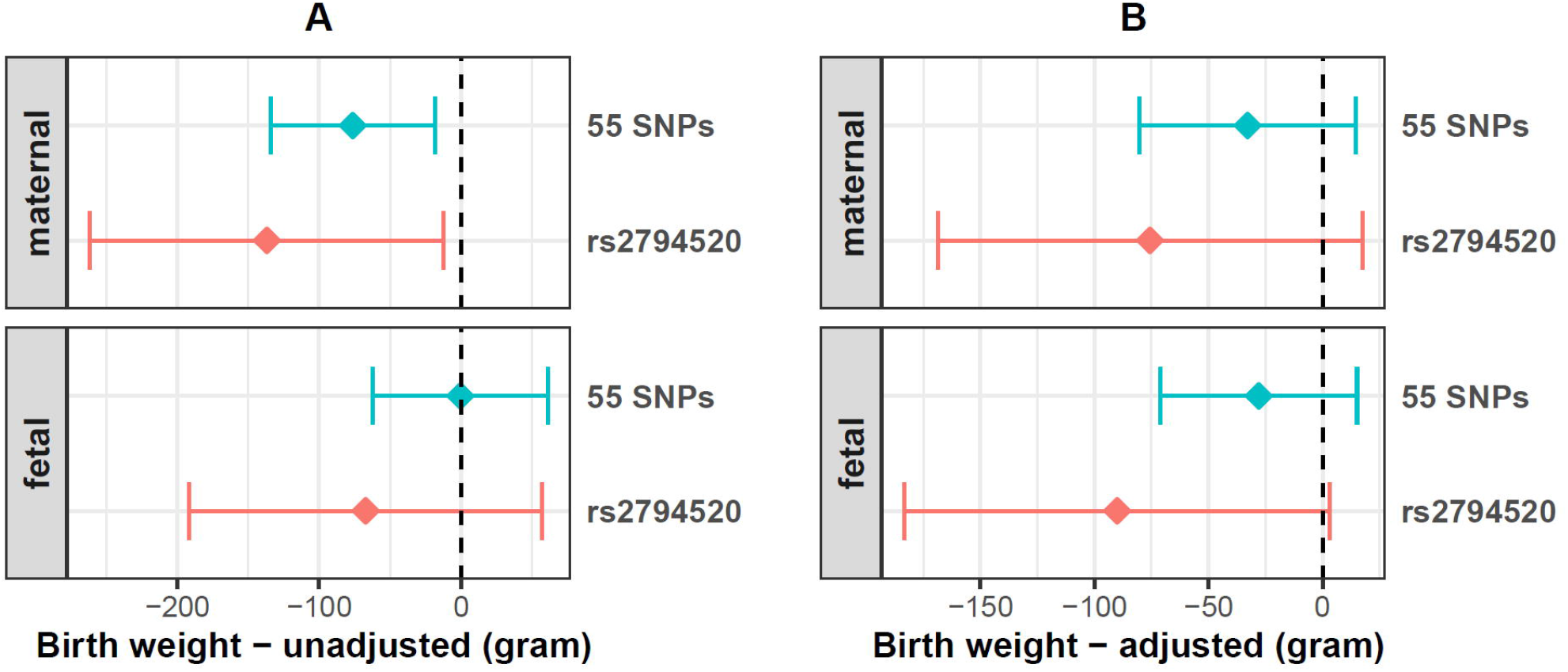
Estimated maternal effects and fetal genetic effects per one unit genetically increased log-transformed CRP levels on unadjusted birth weight (A) and gestational age adjusted birth weight (B). The maternal effects were estimated by maternal non-transmitted alleles (h2) and the fetal genetic effects were estimated by paternal transmitted alleles (h3). The estimates based on multiple (55 SNPs, blue) or single (rs2794520, red) genetic variant(s) are shown.

### Effects of CRP on birth weight estimated using UK Biobank data

We also performed a two-sample MR analysis to estimate the maternal and fetal effects of CRP on birth weight using the UK Biobank GWA summary results for birth weight of the first child (maternal effect) and own birth weight (fetal effect) (available at MR-Base with outcome IDs ‘UKB-a:318’ and ‘UKB-a:198’ respectively). The effect size estimates in the original GWA summary results were reported in standard deviations (SD). For comparison purposes, we converted the unit to grams where 1SD = 668g based on UK Biobank data-field summary (https://biobank.ndph.ox.ac.uk/showcase/field.cgi?id=20022). To account for the maternal and fetal effects of the transmitted alleles in the GWA results, we used the WLM approach [45] to estimate the adjusted maternal and fetal effects (S5 Table). Importantly, these GWA results and effect size estimates of birth weight were not adjusted for gestational duration. Among the 55 CRP-associated SNPs, SNPs with potential horizontal pleiotropic effects identified by MR-PRESSO were removed from the MR analysis (S6 Table).

As shown in Table 3, both unadjusted and WLM-adjusted maternal effects of the CRP associated SNPs on birth weight were negatively correlated with their effect on CRP concentration, indicating a negative causal association. The estimated effect size was −50.1g/log(CRP) (95% CI: −75.5, −24.7, P = 9.6E-5) based on the WLM adjusted maternal effect, i.e., every unit genetically increase in log-transformed maternal CRP level was associated with 50.1g decrease in birth weight of the first child (Figure 4 and S4 Figure). The estimated effect was lower but still significant based on the unadjusted result (birth weight of first child) (effect size = −31.4g/log(CRP), 95% CI: −53.4, −9.4, P = 0.005). The MR analysis using the single IV (rs2794520) showed similar results (Table 3 and Figure 4). Interestingly, we also observed a positive association between the effect on CRP concentration and the WLM adjusted fetal effect on birth weight.

**Table 3.** MR analysis of CRP on birth weight based on UK Biobank GWA summary results.

| Outcome<sup>a</sup> | MR based on 55 CRP SNPs |  |  |  | MR based on rs2794520 |  |  |  |
| --- | --- | --- | --- | --- | --- | --- | --- | --- |
|  | Estimate | Std Err | 95% CI | P-value | Estimate | Std Err | 95% CI | P-value |
| Birth weight of first child | -31.4 | 11.4 | -53.4, -9.4 | 0.005 | -42.8 | 17.4 | -76.2, -8.7 | 0.014 |
| Own birth weight | 10.7 | 7.4 | -3.34, 24.7 | 0.13 | 7.35 | 12.0 | -17.4, 31.4 | 0.57 |
| Birth weight (Maternal effect)<sup>b</sup> | -50.1 | 12.7 | -75.5, -24.7 | 9.59E-05 | -61.5 | 24.7 | -109.6, -13.4 | 0.012 |
| Birth weight (Fetal effect)<sup>b</sup> | 33.4 | 10.7 | 12.7, 54.8 | 0.002 | 37.4 | 20.0 | -2.0, 76.8 | 0.061 |

a: The effect size estimates of birth weight are shown in gram
b: Maternal effect and fetal effect of birth weight were estimated by the weighted linear model (WLM)

**Figure 4.**
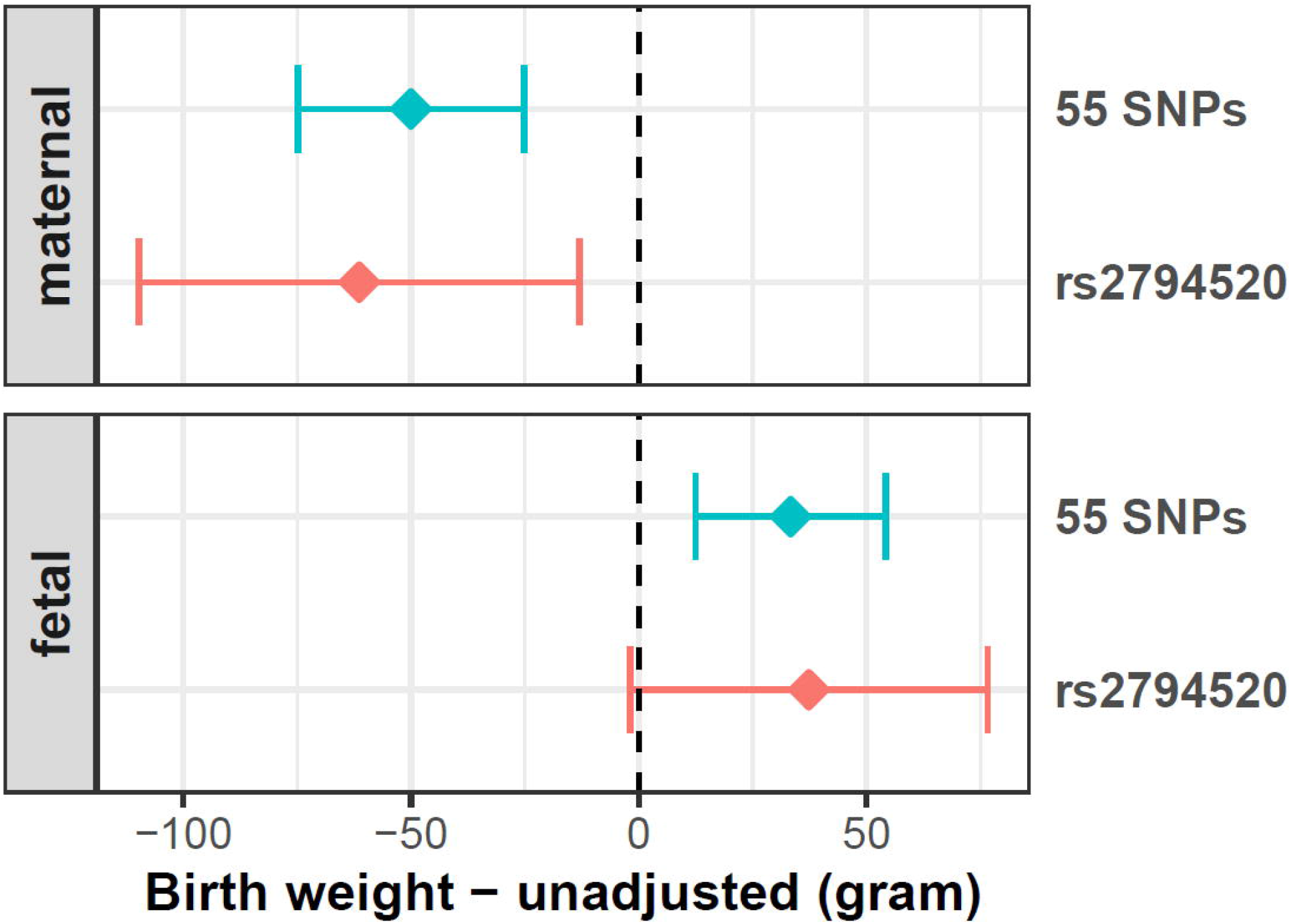
Estimated maternal effect and fetal genetic effect per one unit genetically increased log-transformed CRP levels on unadjusted birth weight using WLM adjusted UKBB summary results. The maternal effect and fetal genetic effect were estimated respectively using WLM adjusted maternal and fetal genetic effects on birth weight (not adjusted by gestational age). The estimates based on multiple (55 SNPs, blue) or single (rs2794520, red) genetic variant(s) are shown.

Because birth weight is influenced by the duration of gestation, we explored whether the observed causal effect of maternal CRP on unadjusted birth weight in the UK Biobank results might be driven by the effect of maternal CRP on gestational duration. Considering that birth weight increases by 25g per day on average [60], we estimated the maternal effect on birth weight that was mediated by gestational duration to be −39.5g/log(CRP) or −68.8g/log(CRP), based on the multiple SNPs (−1.58day/log(CRP)) or the single SNP rs2794520 (−2.75day/log(CRP)) estimates respectively. This effect is comparable with the effect estimated based on the UK Biobank data. Together with the smaller estimated causal effects of maternal CRP on gestational age adjusted birth weight compared to the unadjusted birth weight from the haplotype-based MR analysis in mother/child pairs (Figure 3), these results collaboratively suggest that the causal effect of maternal CRP on birth weight is partially driven by its effect on gestational duration.

## Discussion

We estimated the effects of CRP on gestational duration and birth weight using haplotype-based MR analysis in over 10,000 mother/child pairs. Our results showed that genetically increased maternal CRP was associated with a shorter gestational duration, suggesting an etiological role of maternal CRP on gestational duration. We also found evidence of an effect of higher maternal CRP on lower birth weight, which was in part mediated by the effect of CRP on gestational duration. We observed similar effect estimates of CRP on birth weight using a different (WLM) method to adjust for fetal genotype, in an independent dataset (summary data from UK Biobank), strengthening the credibility of our haplotype-based MR analyses in mother/child pairs. However, we acknowledge that we could not directly estimate the effects of CRP on gestational duration using this alternative method, due to the limited availability of gestational duration data in UK Biobank.

Many epidemiological studies have suggested that elevated maternal serum CRP levels during pregnancy, especially in early pregnancy, are associated with an increased risk of preterm birth [28–31]. The estimated odds ratios are between two to three when using a cut-off of CRP around 8mg/L. One study also showed a dosage effect – every 5mg/L increase in CRP is associated with a 1.4-fold increase in preterm risk [30]. Whilst direct comparisons between our results (which were evaluated by log-transformed CRP) and these studies are difficult, due to the different ways in how CRP and gestational duration were categorized, our findings are broadly consistent with these observational studies. For example, two studies [61,62] reported that a unit change in log-transformed maternal CRP was associated with a reduction of 1.1 and 1.3 gestational days, which is in the same range as our estimated maternal effects based on non-transmitted alleles (Figure 2).

Our analysis also showed negative associations between maternal genetically elevated CRP and birth weight. After adjusting for gestational duration, the magnitude of the association was reduced but not eliminated (Figure 3), indicating a negative impact of maternal CRP on fetal growth independent of its effect on gestational duration. This observation aligns with previous epidemiological reports that elevated maternal CRP levels are associated with small for gestational age or lower birth weight even after adjustment of gestational duration [32,52,62]. We also observed marginal negative associations between paternally transmitted CRP associated alleles (h3) and adjusted birth weight in our haplotype-based analysis (Figure 3), suggesting possible negative fetal genetic effects of CRP increasing alleles on fetal growth. However, this observation is inconsistent with the positive fetal effect on unadjusted birth weight estimated from the UK Biobank data by the WLM method (Figure 4). Further research into the fetal genetic effects of CRP on fetal growth is warranted.

The mechanisms underlying the associations between genetically elevated maternal CRP and shorter gestation and low birth weight need to be further explored. Given the pervasive involvement of inflammation during the whole course of pregnancy, it is reasonable to hypothesize that maternal CRP could influence pregnancy phenotypes at different stages of pregnancy. During early pregnancy a higher CRP level may disturb the normal establishment of pregnancy and increase the risks of adverse outcomes. In late pregnancy, a higher CRP level might reduce fetal growth and trigger the early onset of parturition by stimulating the transition from an anti-inflammatory to a pro-inflammatory state. As an acute-phase protein, CRP has a short half-life (∼19hours) [10], and the genetic variants evaluate an individual ‘s baseline CRP or the potential of CRP production in response to various inflammatory challenges. Therefore, our results may suggest that mothers who are inherently more sensitive to inflammatory stimuli to CRP production are at higher risk of pregnancy complications.

The suggested causal effect of CRP makes it possible to consider CRP not only as a biomarker of shorter gestational duration and lower birth weight, but also as a potential therapeutic target for preventing these pregnancy complications. To date, no trials of specific CRP lowering drugs during pregnancy have been conducted. However, several clinical trials have shown the beneficial effects of low-dose aspirin in preventing preterm birth [63,64] and pregnancy loss [65], especially in women with higher CRP [66]. Pre-clinical studies also showed that statin – a potent agent that can reduce CRP levels independent of its effect on low-density lipoprotein cholesterol [67] – has therapeutic potential in preventing preterm birth [68,69]. Many anti-inflammatory diets or lifestyle modifications that can lower CRP levels have also shown benefits in pregnancy [70].

To our knowledge, this is the first MR study of the potential effects of maternal pregnancy CRP on gestational duration and birth weight. A major advantage of this study is using the haplotype-based approach to explicitly distinguish maternal causal effect and fetal genetic effect. We compared different genetic instruments, MR methods (WLM adjusted two-sample MR using UK Biobank data), and the consistency across results, strengthen confidence in our results. Our study has some limitations. CRP levels during pregnancy were not measured in our study samples which makes it impossible to evaluate the association between genetic instruments and maternal CRP levels. Instead, we used the estimates from a large GWA study of nonpregnant women and men [24], and the effect sizes might differ in pregnant women. We showed similar associations with CRP measured in women only (using ALSPAC data). Also, the consistency of our results with observational studies with CRP measured during pregnancy makes it likely that the genetic instruments are relevant to CRP levels in pregnant women. Our haplotype-based approach requires mother/child pairs, limiting the number of samples. We only observed statistically significant results for the continuously measured traits (e.g., gestational duration and birth weight) rather than the more clinically interesting binary outcome (e.g., preterm birth), possibly due to limited power. Horizontal pleiotropy is always an issue in causal inference using genetic instruments, especially when many genetic variants are used [71]. To mitigate this problem, we used the MR-PRESSO method [56] to detect and remove SNPs with pleiotropic effects. In addition, we used a single SNP (rs2794520), a known protein QTL near the CRP gene as a single genetic instrument that is less prone to bias by pleiotropy [19,20]. It should also be noted that the samples for this study were collected mainly from high-resource European countries, and it is necessary to replicate our findings in diverse populations to investigate whether the observed effects of maternal CRP on gestational duration and birth weight could be modified by genetic background or environmental factors.

In conclusion, we showed that genetically elevated maternal CRP levels are associated with shorter gestational duration and lower birth weight, with the latter partly, though not completely, mediated by the effect on gestational duration. The inferred effect sizes are consistent with previous observational studies. Collectively, these results support the causal role of maternal higher CRP levels on shorter gestational duration and low birth weight. These findings highlight the central role of inflammation in pregnancy and the need for randomized trials to explore the impact of lowering CRP for the prevention of adverse pregnancy outcomes in high-risk women.

## Data Availability

All data produced in the present work are contained in the manuscript

## Acknowledgement

We thank the participants in the Finnish birth cohort as well as the research group who collected the data. We are grateful to all the participating families who took part in the MoBa cohort study. We are grateful to all the families who took part in ALSPAC, the midwives for their help in recruiting them, and the whole ALSPAC team, which includes interviewers, computer and laboratory technicians, clerical workers, research scientists, volunteers, managers, receptionists, and nurses. We are grateful to all DNBC families who took part in the study. We would also like to thank everyone involved in data collection and biological material handling. We would like to acknowledge the participants and research personnel at the participating HAPO field centers. We thank the infants and their parents who agreed to take part in the GPN study and the medical and nursing colleagues who collected that data. We thank dbGAP for depositing and hosting the phenotype and genotype data of the DNBC, HAPO, and GPN data sets.

## Supporting Information

### S1 Text: Supplementary Text (suppl_text.pdf)

### Supplementary Tables (suppl_tables.xlsx)

S1 Table. Descriptive statistics of maternal traits and birth outcomes in the six birth data sets

S2 Table. CRP associated SNPs used as genetic instruments

S3 Table. Haplotype-based association results of the 55 CRP SNPs on gestational duration, preterm birth, unadjusted and adjusted birth weight

S4 Table. MR-PRESSO analysis for haplotype-based MR tests

S5 Table. UK Biobank GWA summary results and WLM estimates of maternal and fetal effects on birth weight

S6 Table. MR-PRESSO analysis for UK Biobank data

### Supplementary Figures (suppl_figures.pdf)

S1 Figure. Scatter plots of MR IVW estimating CRP effects on gestational duration based on maternal transmitted (h1), maternal non-transmitted (h2), and paternal transmitted alleles (h3).

S2 Figure. Scatter plots of MR IVW estimating CRP effects on unadjusted birth weight based on maternal transmitted (h1), maternal non-transmitted (h2), and paternal transmitted alleles (h3).

S3 Figure. Scatter plots of MR IVW estimating CRP effects on adjusted birth weight based on maternal transmitted (h1), maternal non-transmitted (h2), and paternal transmitted alleles (h3).

S4 Figure. Scatter plots of MR IVW estimating CRP effects on birth weight based on WLM-adjusted maternal (left) and fetal effects (right) estimated from UKBB data.

