## Supplementary Text for "Interrogating the causal effects of maternal circulating CRP on gestational duration and birth weight"

### Data sets

We used phenotype and genomewide SNP data of 10,734 mother/infant pairs from six birth studies collected from UK, Northern Europe, Australia, and North America.

#### *FIN*

The Finnish dataset (FIN) was collected for a genetic study of spontaneous preterm birth [1]. Briefly, whole blood samples were collected from more than 1,600 mother/child pairs from the Helsinki (southern Finland) University Hospitals between 2004 and 2014. All the studied samples are of Finnish descent. Crown-rump length at the first ultrasound screening between 10+ and 13 weeks was used to determine the gestational age. 2,962 blood samples from mothers and children were genotyped. After genotype quality control (QC) procedure and applying the phenotype-based inclusion/exclusion criteria, 1,170 mother/child pairs were selected and used in the analysis. The study was approved by the Ethics Committee of Oulu University Hospital and that of Helsinki University Central Hospital. Written informed consent was given by all participants.

#### *MoBa*

The Mother Child dataset of Norway (MoBa) is a nationwide Norwegian pregnancy study administered by the Norwegian Institute of Public Health. The study includes more than 114,000 children, 95,000 mothers and 75,000 fathers recruited from 1999 through 2008 [2]. Gestational age was estimated by ultrasound at gestational weeks 17–19. In the few cases without ultrasound dating, gestational age was estimated using the date of the last menstrual period. For the current study, we used the mother-child pairs that were selected from Version 4 of the MoBa dataset, which included a total of 71,669 pregnancies [3]. Singleton live-born spontaneous pregnancies with mothers in the age group 20–34 years were selected. Random sampling was done from two gestational age ranges 154-258 days (cases) and 273–286 days (controls). Pregnancies involving pre-existing medical conditions, pregnancies with complications as well as pregnancies conceived by in vitro fertilization, were excluded from the study. In total, blood samples from 3,120 mothers and children were genotyped [3]. 1,009 mother/child pairs that passed QC and inclusion/exclusion criteria were included in the analysis. All parents gave informed, written consent. The study was approved by The Regional Committee for Medical Research Ethics in South-Eastern, Norway.

*DNBC*

The Danish National Birth Cohort (DNBC) followed over 100,000 pregnancies between 1996 and 2003 with extensive epidemiologic data on health outcomes in both mother and child [4]. The current study used the data downloaded from the Database of Genotypes and Phenotypes (dbGaP) (phs000103.v1.p1), which contains data from a genome-wide case/control study using approximately 1,000 preterm mother-child pairs (gestational age between 22–37 weeks) from the DNBC, along with 1,000 control pairs in which the child was born at ~40 weeks gestation. Gestational duration in this dataset was determined by a consensus algorithm combining all available information from multiple sources: self-reported date of last menstrual period, self-reported delivery date, and gestational age at birth registered in the Medical Birth Register and the National Patient Register. Of the 3,886 samples with genotype data, we identified 1,739 mother/child pairs and used them in the analysis. The study protocol was approved by the Danish Scientific Ethical Committee and the Danish Data Protection Agency.

*HAPO*

The Hyperglycemia and Adverse Pregnancy Outcome (HAPO) Study [5,6] is a multicenter, international study in which high quality phenotypic data related to fetal growth and maternal glucose metabolism has been collected from 25,000 pregnant women of varied racial and socio-demographic backgrounds using standardized protocols that were uniform across centers. For the current study, we utilized phenotype and genotype data of 1,500 infants and their mothers of European descent download from dbGaP (phs000096.v2.p1). The samples were collected from Toronto, Canada, Belfast, UK, Brisbane and Newcastle, Australia. Gestational duration in this dataset was determined by last menstrual period or ultrasound estimation from 6-24 weeks. Of the 2,866 samples with genotype data, we identified 1,089 mother/child pairs and used them in the analysis.

*GPN*

The Genomic and Proteomic Network for Preterm Birth Research (GPN) study [7] is a multicenter observational genome-wide association study (GWAS) designed to determine the genetic predisposition to idiopathic preterm birth. Phenotype data and genotype data from 743 spontaneous preterm births (20 to less than 34 weeks gestation), and 752 controls (39 to less than 42 weeks gestation) of diverse ethnic background (White, Hispanics, African Americans, and Others) were collected. For this current study, we identified 343 mother/child pairs of European descent from the data downloaded from dbGaP (phs000714.v1.p1).

#### *ALSPAC*

The Avon Longitudinal Study of Parents and Children (ALSPAC) is a prospective birth cohort study. 14,541 pregnant women resident in the former county of Avon (situated around the city of Bristol in the South West of England) with expected dates of delivery 1st April 1991 to 31st December 1992 were recruited [8,9]. The children arising from these women, and their partners were followed up intensively over nearly three decades. Genotype data of the mothers and children were generated using the Illumina HumanHap550 quad (children) and Illumina human660W quad (mothers). This resulted in a dataset of 17,842 participants (either mothers or offspring), containing 6,305 mother-offspring pairs, each with 465,740 SNPs genotyped. From this data set, 5,384 mother-offspring pairs who passed genotype QC and inclusion/exclusion criteria were included in the analysis. Please note that the study website contains details of all the data that is available through a fully searchable data dictionary and variable search tool (http://www.bristol.ac.uk/alspac/researchers/our-data/). Ethical approval for the study was obtained from the ALSPAC Ethics and Law Committee and the Local Research Ethics Committees.

### Phenotype data

#### *Phenotype measures*

In this study, we focused pregnancy outcomes including gestational duration (and preterm birth as a binary trait) and birth weight. Maternal height, weight and most of the birth outcomes were available in most of the data sets (except for birth weight and length were not available from MoBa and birth length was not available in the DNBC data set). In the ALSPAC study, serum CRP concentration was measured in some mothers in a follow-up data collection 19 years after the pregnancy [8,9].

#### *Inclusion/exclusion criteria*

We studied singleton pregnancies with spontaneous live births – including spontaneous deliveries with or without premature rupture of membranes (PROM). C-sections after spontaneous onset of labor were retained. Medically indicated deliveries or C-sections and repeat C-sections were excluded. Mother/child pairs without gestational duration information were excluded. Pregnancies with known gestational or fetal complications (e.g. placental abnormities, chorioamnionitis, preeclampsia, and congenital anomalies) and pregnancies involving pre-existing medical conditions (i.e. hypertension or diabetes) or maternal risk exposure (e.g. drug use during pregnancy) known be associated with preterm birth were also excluded. Pregnancies with gestational diabetes and gestational hypertension were not excluded for the investigation of maternal physiological changes during pregnancy.

### Genotype data

The genomewide SNP data of the data sets were generated using either Affymetrix 6.0 or various Illumina arrays. After genotype calling, similar genotype QC procedures were applied.

#### *Genotype calls*

For the FIN dataset, genotyping was conducted using Affymetrix 6.0 (Affymetrix, California, United States) and various other Illumina arrays (Illumina, California, United States). For the Affymetrix SNP Array 6.0, genotype calls were determined using the CRLMM algorithm [10,11] among chips that passed the vendor-suggested QC (Contrast QC > 0.4). For the Illumina chips, the genotype calling was conducted using Illumina's genotyping module v1.94 in the GenomeStudio v2011.1.

The samples from the MoBa dataset were genotyped using the Illumina Human660W-Quadv1_A bead chip (Illumina Inc.) and the genotype calls were determined using CRLMM algorithm.

The DNBC samples were genotyped using Human660W-Quad bead arrays from Illumina. The raw genotype intensity data (*.idat) files were obtained from dbGaP and we performed genotype calls using CRLMM algorithm.

The HAPO samples of European descent were genotyped using Human610-Quad array. We obtained the raw genotype intensity data (*.idat) files from dbGaP and performed genotype calls using CRLMM algorithm.

The processed genotype calls in plink format of the GPN data set was obtained from dbGaP (phs000714.v1.p1). Data from participants of apparent duplications with others (IBD>0.8), with sex discrepancies (between known sex and genetically inferred sex), and children with high Mendelian errors (>10%) were deleted.

The ALSPAC genotype data were generated using the Illumina HumanHap550 quad (children) and Illumina human660W quad (mothers). The cleaned genotype calls of 465,740 SNPs of 17,842 individuals were obtained from ALSPAC.

*Genotype QC procedures*

We performed similar genotype QC across of all the dataset. We first performed sample-level QC based on call rate, overall heterogeneity, sex discrepancies; we checked the pedigree relationship based IBD analysis; Genetic ancestry was assessed by principal components analysis (PCA) anchored by 1000 Genomes reference samples. Individuals with non-European ancestry were excluded. We then perform marker level QC: SNPs with low call rate (<98%), low minor allele frequency (<0.01) or significant deviation from Hardy-Weinberg Equilibrium ($p < 5\times10^{-6}$) were excluded.
