## Supplementary Figures for "Interrogating the causal effects of maternal circulating CRP on gestational duration and birth weight"

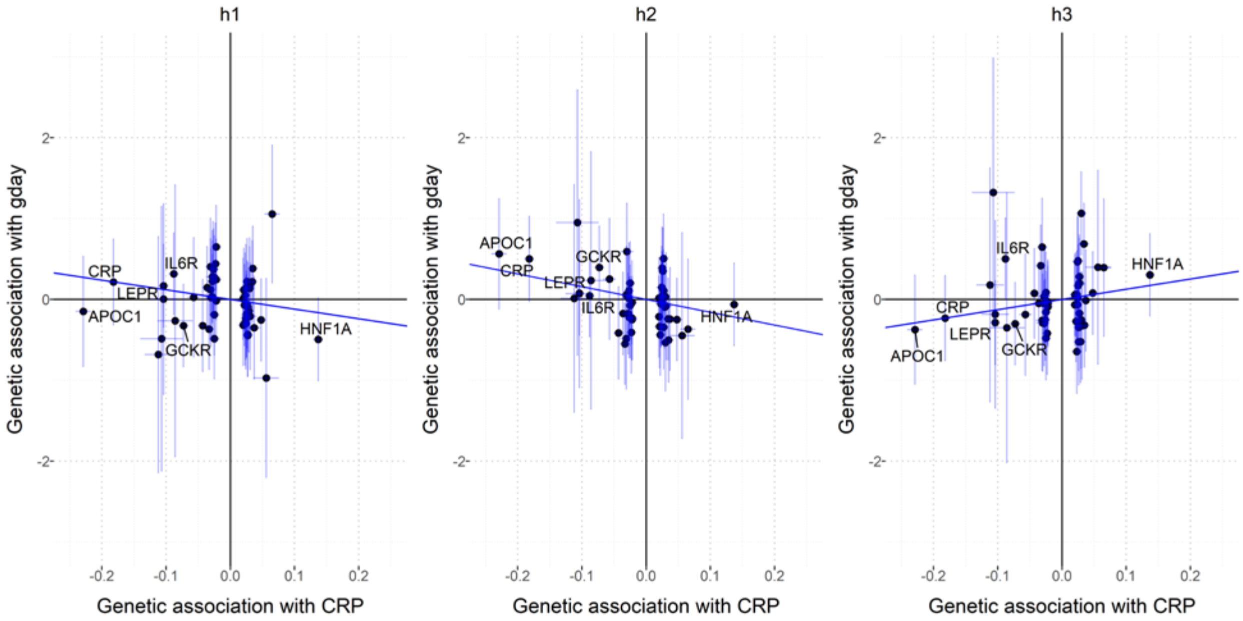


S1 Figure. Scatter plots of MR IVW estimating CRP effects on gestational duration based on maternal transmitted (h1), maternal non-transmitted (h2), and paternal transmitted alleles (h3).

The effect size estimate of gestational duration is shown in days. The labels are the genes of the top 6 CRP IVs with association P < 1E-50. The fitted IVW line is colored blue.

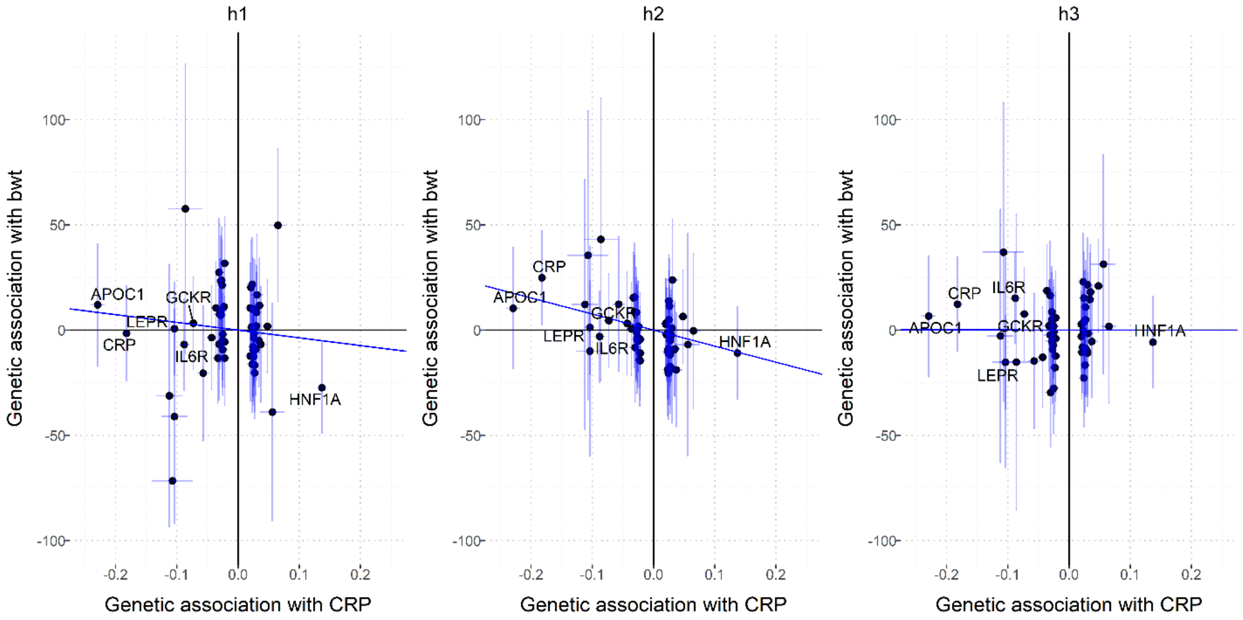


S2 Figure. Scatter plots of MR IVW estimating CRP effects on unadjusted birth weight based on maternal transmitted (h1), maternal non-transmitted (h2), and paternal transmitted alleles (h3).

The effect size estimate of gestational duration is shown in days. The labels are the genes of the top 6 CRP IVs with association P < 1E-50. The fitted IVW line is colored blue.

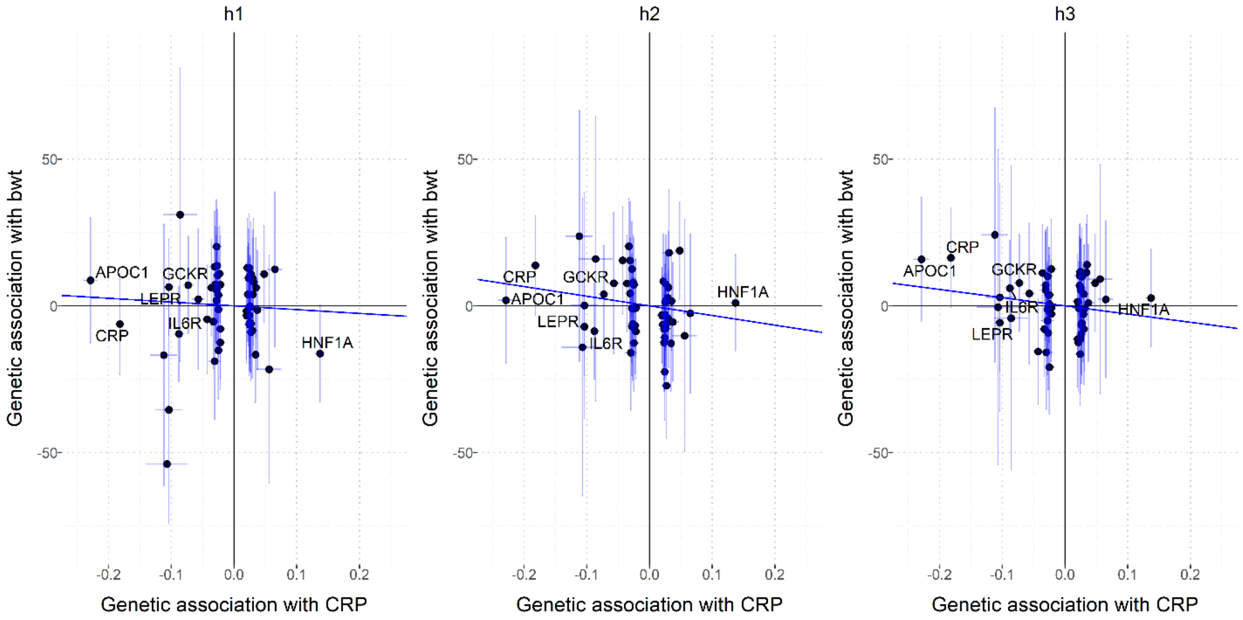


S3 Figure. Scatter plots of MR IVW estimating CRP effects on adjusted birth weight based on maternal transmitted (h1), maternal non-transmitted (h2), and paternal transmitted alleles (h3).

The effect size estimate of gestational duration is shown in days. The labels are the genes of the top 6 CRP IVs with association P < 1E-50. The fitted IVW line is colored blue.

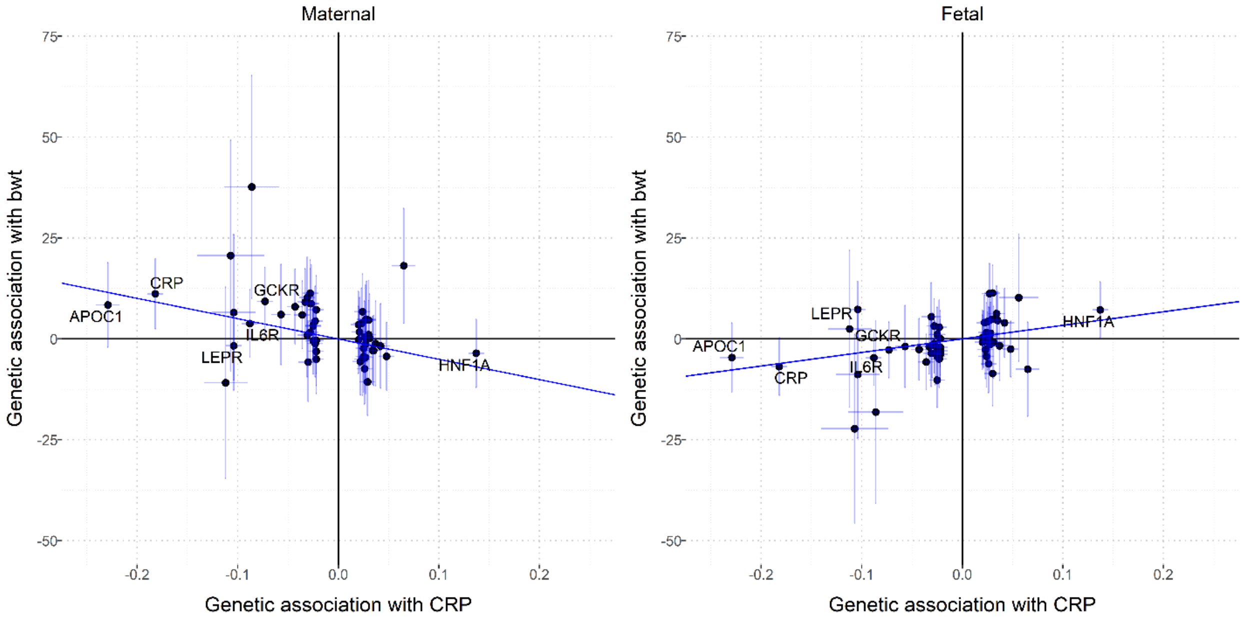


### S4 Figure. Scatter plots of MR IVW estimating CRP effects on birth weight based on WLM-adjusted maternal (left) and fetal effects (right) estimated from UKBB data.

The effect size estimate of birth weight is shown in standard deviations (SD). The labels are the genes of the top 6 CRP SNPs with association P < 1E-50. The fitted IVW line is colored blue.
